# Network-based Molecular Constraints on *in vivo* Synaptic Density Alterations in Schizophrenia

**DOI:** 10.1101/2025.03.22.25324465

**Authors:** Sidhant Chopra, Patrick D. Worhunsky, Mika Naganawa, Xi-Han Zhang, Ashlea Segal, Loïc Labache, Edwina Orchard, Vanessa Cropley, Stephen Wood, Gustavo A. Angarita, Kelly Cosgrove, David Matuskey, Nabeel B. Nabulsi, Yiyun Huang, Richard E. Carson, Irina Esterlis, Patrick D. Skosnik, Deepak C. D’Souza, Avram J. Holmes, Rajiv Radhakrishnan

## Abstract

Converging neuroimaging, genetic, and post-mortem evidence highlights the fundamental role of synaptic density reductions in schizophrenia pathogenesis. However, the brain-wide spatial pattern of these alterations and the mechanisms underlying this patterning remain to be established. Here, using [^11^C]UCB-J radiotracer positron emission tomography (PET) imaging in individuals with schizophrenia (n=29) and healthy controls (n=93), we find a prominent and widespread pattern of lower synaptic density (0.58 < Cohen’s *D* < 1.47; p_FWE_<0.05) in patients. The left hemisphere is substantially more impacted than the right (Cohen’s *D* = 1.14; p < .001), with frontal, temporal, cingulate, thalamic, striatal and hippocampal areas particularly affected. Synaptic density alterations were not spatially aligned with gray matter alterations indexed using anatomical Magnetic Resonance Imaging. Lower synaptic density in the left hemisphere is associated with higher normative concentrations of GABA_A/BZ_, 5HT_2A_, mGluR5 and 5HT_1B_ (𝑟_𝑐𝑐𝑎_ = .68; 𝑝 = .022). Simulation-based network diffusion models identified regions that may represent the initial sources of pathology, nominating left inferior frontal areas (𝑝_𝐹𝖶𝐸_ < .05) as potential foci from which synaptic pathology initiates and then propagates to structurally connected and molecularly similar areas. Overall, our findings provide *in vivo* evidence for widespread synaptic density deficits in schizophrenia that are left-lateralized, independent of gray matter alterations, aligned to specific neurochemical systems, and suggest that such synaptic pathology may propagate in a pattern consistent with axonal networks.

## Introduction

Post-mortem, genetic and neuroimaging studies have converged to implicate loss of synapses as a fundamental pathophysiological process in schizophrenia^1–4^. The advent of positron emission tomography (PET) radioligands with high specificity for synaptic vesicle proteins^5^ have made it possible to examine presynaptic synaptic terminal density *in vivo*^6^, with preliminary studies in schizophrenia^7–9^ demonstrating reductions in specific cortical and subcortical areas. However, the brain-wide spatial patterning of synaptic pathology, and the mechanisms shaping this pattern, remain poorly understood.

Research in neurological conditions shows that the brains vulnerability to pathology is not spatially random but instead reflects intrinsic properties of large-scale systems^10^, cellular composition^11^, and receptor distributions^12^. Similar principles may apply in schizophrenia, where gray matter morphological deficits preferentially impact higher-order association networks and align with normative receptor expression profiles. Moreover, several lines of evidence suggest that brain pathology in schizophrenia is constrained by macroscale axonal networks. For instance, grey matter reductions in patients correlate with the microstructure of adjacent white matter^13^, across spatially distributed regions^14,15^, with normative connectome organization^16,17^, and with reductions in structurally connected and functionally coupled regions^18–20^. Moreover, multiple longitudinal and modelling-based studies have now demonstrated that pathology may spread from an initial site to axonally connected territories^20–23^. However, widely used MRI-based morphometric measures predominantly index a composite of tissue properties such as intracortical myelin, neurite packing, glial density, iron and water content and vascular effects, rather than synaptic terminals. Thus, direct *in vivo* measures are required to test whether the systems-level features confer vulnerability and shape the spatial organization of synaptic pathology in schizophrenia.

Here, we address this gap by mapping the *in vivo* brain-wide spatial pattern of synaptic density alterations using [^11^C]UCB-J PET in a large single-site cohort of 122 individuals, including 29 with schizophrenia. We demonstrate a prominent, widespread pattern of reduced presynaptic terminal density in schizophrenia, with stronger effects in the left hemisphere and in frontal, temporal, cingulate, thalamic, striatal, and hippocampal regions. These reductions were spatially independent of grey matter volume alterations measured with structural MRI and closely aligned with normative distributions of specific neurotransmitter and transporter systems. Network diffusion models further nominated left prefrontal regions as potential sources of pathology, from which synaptic deficits may propagate to axonally connected and molecularly similar territories. Together, these findings provide the first i*n vivo* evidence that presynaptic terminal deficits in schizophrenia are widespread, left-lateralized, and shaped by both molecular architecture and macroscale network organization.

## Methods

### Sample characteristics

A total of 122 individuals, including 29 individuals diagnosed with schizophrenia were included in this study. Individuals with schizophrenia were recruited via clinician referrals, paper, and web advertisements. The majority of individuals with schizophrenia were recruited using criteria that excluded based on the following criteria: regular exposure to drugs of abuse (except nicotine and caffeine) within the past 3 months based on history or positive urine drug screen, lifetime substance use disorder (except for nicotine and caffeine), weekly alcohol consumption exceeding National Institute on Alcohol Abuse and Alcoholism (NIAAA) guidelines (4 or more drinks on any single day and/or 21 or more drinks per week for males), history of significant head injury resulting in unconsciousness, unstable medical condition, significant prior exposure to radiation, metal in the body which would exclude MRI scan or claustrophobia. A subset of individuals with schizophrenia were recruited using a less restrictive criteria, where substance use was not an exclusion. We repeated our primary analyses after excluding subjects with a history of, or current, substance abuse (n=7; SFig1). Data from 13 of the included individuals with schizophrenia have been part of a previously published study^7^. The healthy control subjects were included from studies performed at the Yale PET Center using the [^11^C]UCB-J PET to investigate brain synaptic density across a variety of neuropsychiatric conditions. Healthy control individuals were excluded for a current and/or lifetime diagnosis of a psychiatric disorder, current or past serious medical or neurological illness (including a history of head injury with loss of consciousness), metal in body which would result in MRI contraindication, or a history of substance abuse or dependence. Demographic and sample characteristics are provided in Table 1. All study protocols were approved by the Yale University Human Investigation Committee and Radiation Safety Committee. All participants provided written informed consent prior to participation.

**Table 1.** Sample Demographics.

|  | Schizophrenia<br>(N=29) | Control (N=93) | p-value |
| --- | --- | --- | --- |
| Age, mean years (SD) | 43.21 (14.66) | 45.40 (16.83) | 0.501 |
| Females, N (%) | 4 (13.8%) | 37 (39.8%) | 0.018 |
| Mass dose, mean in ug/kg (SD) | 0.02 (0.02) | 0.02 (0.02) | 0.647 |
| Activity at time of injection, mean MBq (SD) | 235.53 (201.00) | 194.61 (125.28) | 0.332 |
| Antipsychotic exposure, mean CPZ eqs. (SD) | 2693.34 (2738.91) | - | - |
| PANSS Total, mean (SD) | 68.39 (13.51) | - | - |
| PANSS Positive, mean (SD) | 17.46 (5.26) | - | - |
| PANSS Negative, mean (SD) | 16.64 (5.06) | - | - |
| PANSS General, mean (SD) | 33.93 (6.88) | - | - |

CPZ eqs. = lifetime exposure to antipsychotics as chlorpromazine (CPZ) equivalents (eqs.). Listed p-values correspond to two-sample t-tests, except for gender-differences, where they correspond to a $X^2$ proportion test. PANSS= Positive and Negative Syndrome Scale.

### PET and MR imaging acquisition and processing

[^11^C]UCB-J was synthesized using established methods^24^. All [^11^C]UCB-J PET measurements were conducted on the HRRT (Siemens Medical Solutions), which acquires 207 slices (1.2-mm slice separation) with a reconstructed image resolution of ∼3 mm. Before every [^11^C]UCB-J injection, a transmission scan was performed for attenuation correction. Dynamic PET data (frames: 6×0.5 min, 3×1 min, 2×2 min, and 16×5 min) were acquired and reconstructed using the MOLAR algorithm^25^. Event-by-event motion correction was performed using Polaris Vicra optical tracking (NDI Systems, Waterloo, Canada) with reflectors mounted on the subject’s head via a swim cap^26^. All gray matter voxels were corrected for partial volume effects using the Müller-Gärtner algorithm^27^. Then, the simplified reference tissue model 2 (SRTM2)^28,29^ with the centrum semiovale as the reference region, was applied using 60 mins of dynamic data to generate parametric images of binding potential (*BP*_ND_) ^6,7,30,31^. Mean voxel-level *BP*_ND_ maps for patients and controls are provided in SFig. 2.

A high-resolution T1-weighted structural MPRAGE scan (TR/ TE = 2530/3.34, flip angle = 7°, in-plane resolution = 0.98 × 0.98 mm, matrix size = 256 × 256, slice thickness = 1 mm, slices = 176) was acquired on a 3T MAGNETOM Prisma scanner (Siemens, Erlangen, Germany). Registration of parametric PET images to Montreal Neurological Institute (MNI152) standard space was performed using SPM12 (Wellcome Trust center for Neuroimaging, London, UK)^32^. For each subject, PET images were motion-corrected by registering each frame to a summed image (0–10 min post-injection). A linear registration aligned the summed PET image to the T1-weighted MR image, followed by a nonlinear registration to the MNI template. The combined transformations were then applied to the parametric PET image. The standardized parametric PET images were smoothed with a 4mm full width half maximum Gaussian kernel then masked using an enhanced gray-matter tissue prior map thresholded at .25^33^.

### Mapping brain-wide synaptic density alterations

General liner models examining lower *BP*_ND_ in individuals with schizophrenia compared to controls, adjusting for age and sex, were fit at each grey matter voxel. Non-parametric voxel-level family-wise error (FWE) correction was implemented using CAT12^34^, using 5000 permutations, with significance assessed at p_FWE_ < 0.05 (Fig 1A).

**Fig. 1.**
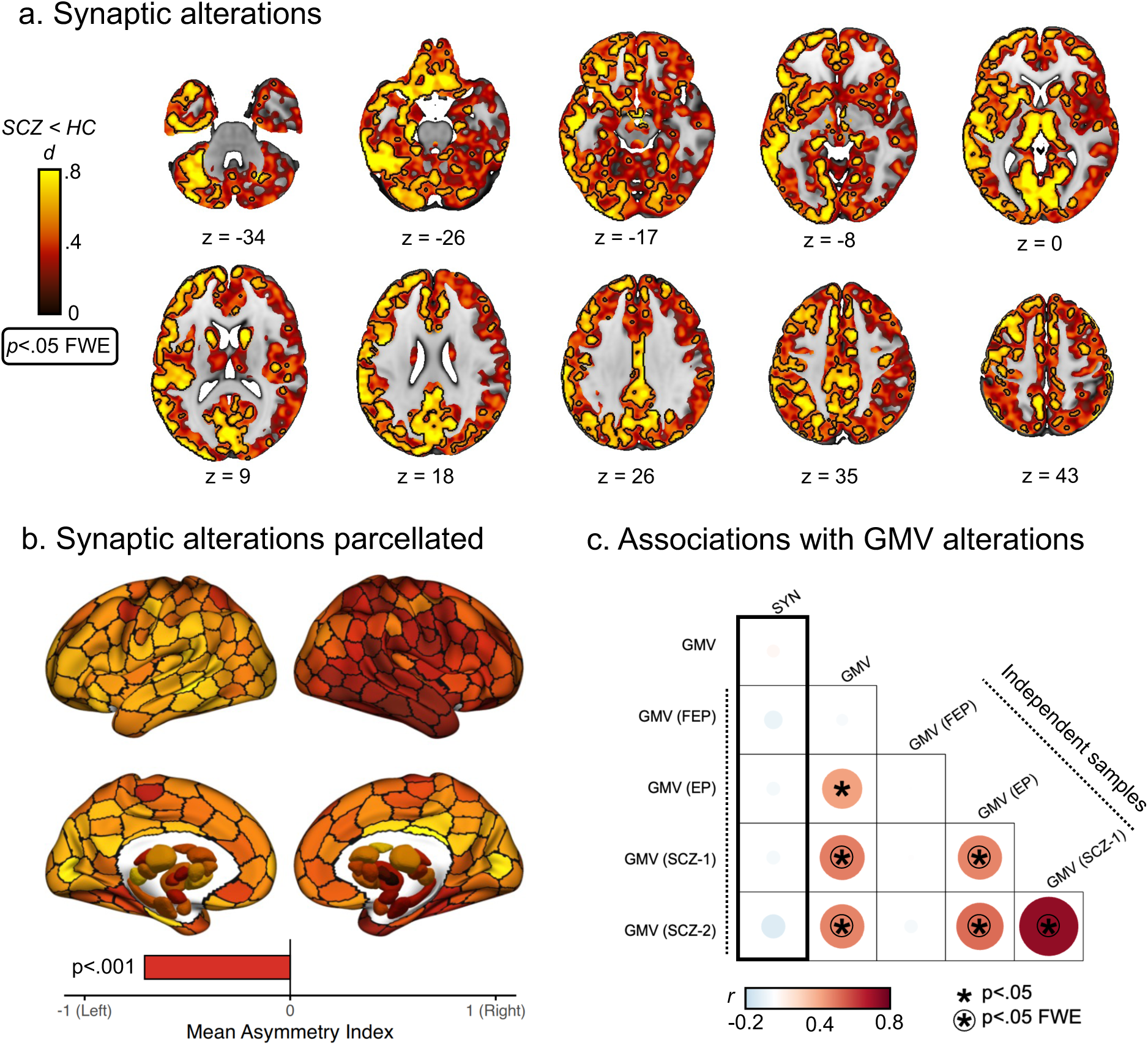
Widespread and left-lateralized lower synaptic density in schizophrenia. **a)** Voxel-level effect-size map of difference in binding potential (non-displaceable) between individuals with schizophrenia (SCZ) and healthy controls (HC), adjusting for age and sex. Test statistics (t-values) were converted to standardized effect-sizes (Cohen’s *D*), with the black outline indicated voxel-level family-wise error corrected (FWE) significance at p<0.05. **b)** Effect-size map parcellated using the 300 homotopic cortical regions^35^ and subcortical Tian 32 region^36^ atlases, with the mean value of voxels assigned to each region. Bottom bar displays mean asymmetry index across all regions, with the p-value representing overall significant leftward laterality in synaptic loss. **c)** Thick black outline highlights spatial correlation (Pearson’s r) between parcellated synaptic density alterations (SYN) and gray matter volume (GMV) within the current sample (top left box), as well as four independent samples including antipsychotic-naive first-episode psychosis (FEP), early psychosis (EP) and two established schizophrenia (SCZ) cohorts. No association between synaptic density alterations and gray matter alterations were found. All other boxes represent correlations between GMV maps between the current and independent samples. Asterisks (*) indicate statistical significance (𝑝 < .05) completed to spatial autocorrelation preserving null models, with a circle indicating survival against FWE correction.

To examine hemispheric asymmetry in brain-wide synaptic pathology, t-statistic map which indexed differences in synaptic density between patients and controls, were then parcellated using a functional previously validated homotopic atlas into 300 discrete cortical^35^ and 32 subcortical^36^ areas of approximately equal size. Hemispheric asymmetry was quantified by computing right-left differences in the test-statistic across each homotopic parcel (negative values indicating leftward asymmetry). A one-sample t-test was used to test for systematic asymmetry. To generate a non-parametric null distribution, left and right values were randomly swapped within each homotopic pair (10,000 permutations), and significance was assessed as the proportion of permuted mean differences exceeding the observed mean difference.

### Association between grey-matter volume and synaptic density alterations

Voxel-based morphometry (VBM), implemented in the CAT12 toolbox^34^, was used to index brain-wise grey matter volume alteration between individuals with schizophrenia and controls within the current sample, as well as four previously published independent samples^20^ which included individuals with medication-naive first-episode psychosis^37^ (N_case/control_=59/27), early psychosis^38^ (N_case/control_=121/57), and two cohorts of established schizophrenia^39,40^ (N_case/control_=66/72; N_case/control_=70/62). The VBM procedure used for each sample has been previously described in detail^20^. The resulting brain-wide voxel-level t-statistic map from each study, which indexed differences in gray matter volume between patients and controls, were then parcellated using the same 332 region atlas described above. The volume alteration of a region was estimated as the mean test-statistic (comparing patients and controls) of all voxels corresponding to that region. The same parcellation procedure was followed for synaptic density alteration map from the current sample (Fig 1B), allowing for direct comparison between synaptic and gray-matter volume alteration maps. Product-moment correlations (r) were used to examine spatial associations between synaptic density alteration and gray matter alterations within the current sample and four independent cohorts (Fig 1C). Statistical significance of associations between spatial maps was assessed using a benchmark null model that preserved spatial autocorrelation, allowing us to evaluate whether the observed findings were specific to the empirically observed alteration maps or were a generic property of the intrinsic spatial structure of these maps. The procedure used a spin-test to rotate cortical region-level t-values from the synaptic density alteration map 10,000 times^41,42^. For brain-wide analyses, the rotation was applied to one hemisphere and then mirrored for the other hemisphere. For hemisphere-specific analyses, the rotations were computed independently for each hemisphere. Sub-cortical t-values were randomly permuted without replacement. All 𝑝-values were quantified as the fraction of null correlation values exceeding the observed correlation, considering both positive and negative tails of the null distribution (two-tailed). Statistical significance was assessed at p<.05 and inference was FWE corrected using Bonferroni correction (Fig 1C).

### Quantifying network, neurochemical and cellular enrichment of *synaptic density alterations*

#### Network-level enrichment

To examine whether synaptic density alterations were preferentially enriched within specific large-scale canonical functional networks and cytoarchitectonic territories of each hemisphere, we used two brain system classifications: i) intrinsic functional networks based on the 17 cortical Yeo^43,44^ and ii) cytoarchitectonic types of laminar differentiation based on the canonical von Economo and Koskinas atlas ^41,45,46^. Synaptic pathology was estimated averaging the t-statistic across regions belonging to each functional network or cytoarchitectonic subdivision (Fig. 2A-B). Statistical significance of observed systems-level synaptic pathology was compared to mean values derived using the spin-test with 10,000 permutations, as previously described. Statistical inference was assessed at p<.05 (two-tailed) and FWE corrected using Bonferroni correction across 17 networks. To quantify the effect size for each network, we computed z-scores by comparing the observed test-statistic to the null distribution^18^.

**Fig. 2.**
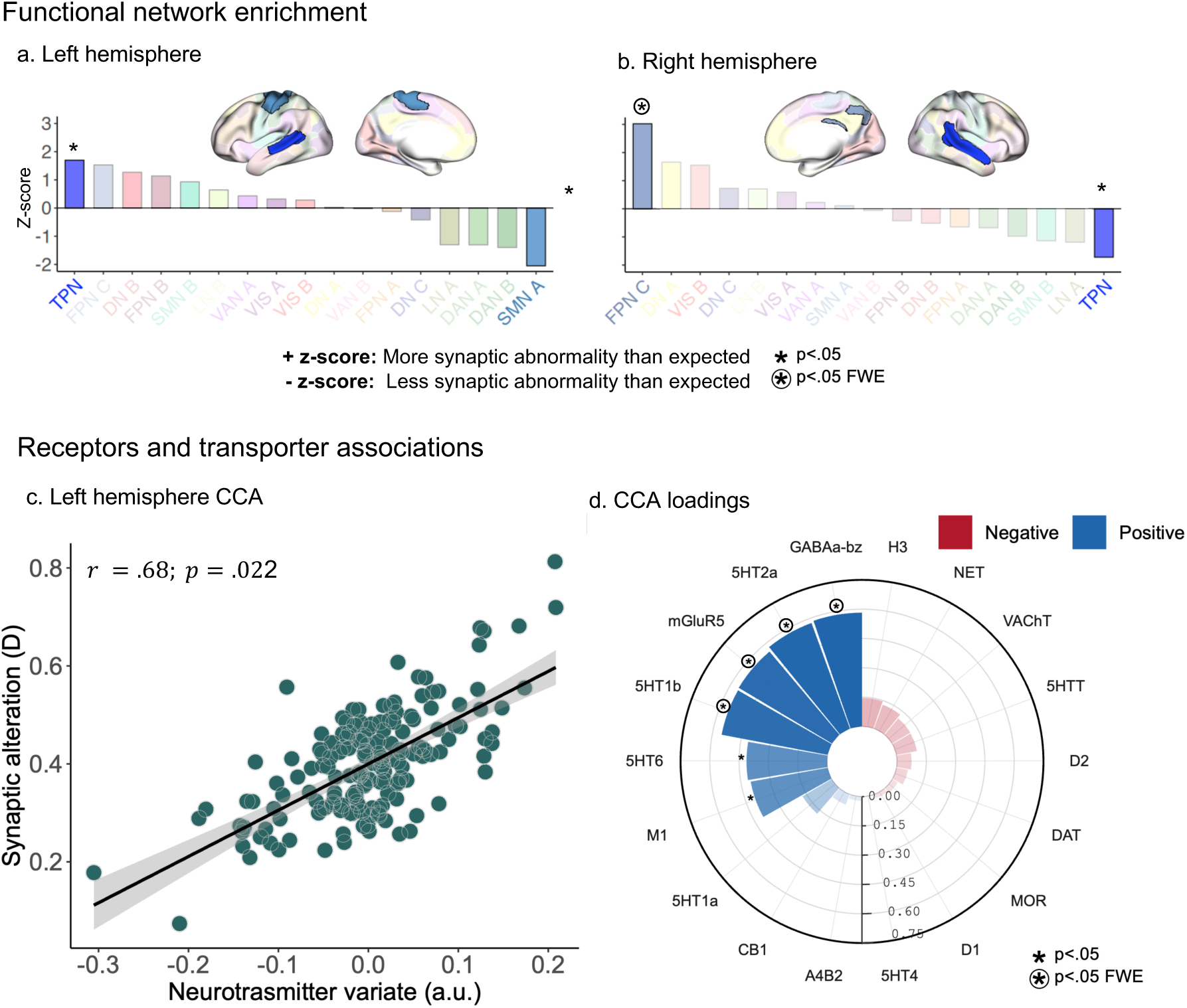
Network and neurochemical correlates of synaptic pathology in schizophrenia. **a-b)** Enrichment of synaptic pathology within left (a) and right (b) hemisphere functional networks. Bars plot each networks corresponding z-scores relative to the null distribution generated by the spatial permuting procedure. A positive z score indicates greater synaptic loss than expected, whereas a negative z-score indicates less synaptic loss than expected. Asterisks (*) indicate statistical significance (𝑝 < 0.05), with a circle indicating survival against family-wise error (FWE) correction. **c)** Significant canonical correlation between regional synaptic deficits and normative receptor/transporter density in the left hemisphere. The synaptic variate (y-axis) in this case is the regional synaptic deficit/effect size, whereas the neurotransmitter variate is a normalized weighted combination of regional receptor/transporter densities. The non-parametric *p*-value was computed using the spatial permuting procedure. **d)** Canonical loading (Pearson correlation between normative receptor/transporter density data and neurotransmitter variate) corresponding to (c), where the loading of each receptor/transporter map onto the significant neurochemical variate are plotted. positive loadings indicate that lower synaptic density is associated with higher levels, whereas negative loadings indicate that lower synaptic density is associated with lower levels of the specific molecular system. Asterisks (*) indicate statistical significance using bootstrapping (p<0.05), with a circle indicating survival of FWE correction. TPN: temporoparietal network, DN: default network, FPN: frontoparietal network (sometimes referred to as cognitive control network), LN: limbic network, VAN: salience/ventral attention network, DAN: dorsal attention network, SMN: somatomotor network, VIS: visual network.

### Neurochemical and cellular associations

To examine whether synaptic density alterations were preferentially enriched with normative neurochemical or cellular densities, we used two sources of data. For regional normative neurochemical densities, we used 18 different group-consensus PET binding maps from healthy population collated by the *neuromaps* toolbox^47^. Following previous work, for tracers that had multiple samples, a sample-size weighted average map was computed^48^. For cases where two different tracers targeted the same receptor/transporter, the one with the larger sample size was retained. Further details regarding each PET map are provided elsewhere^47^. Each PET map was masked and parcellated into the same 332 region atlas as the synaptic density alteration maps. Regional transcriptomic cell-type concentrations were deconvolved from whole brain bulk tissue transcriptions provided by the Allens Human Brain Atlas (AHBA)^49^, with 24 different cell-types being imputed using recently published single-nucleus RNA-sequencing data^50^ (see Zhang et al^51^ for detailed overview). Briefly, the transcription signatures identifying each class of cell in the AHBA bulk samples were derived from cortical single-nucleus RNA sequencing (snRNA-seq) data of eight cortical areas. This included 24 cellular classes with distinct laminar specialization, developmental origin, morphology, spiking pattern, and broad projection targets^52^. These cells include 9 GABAergic inhibitory interneurons (PAX6, SNCG, VIP, LAMP5, LAMP5 LHX6, Chandelier, PVALB, SST CHODL, SST), 9 glutamatergic excitatory neurons (L2/3 IT, L4 IT, L5 IT, L6 IT, L5 ET, L5/6 NP, L6 CT, L6b, L6 IT Car3), and 6 non-neuronal cells (Astro, Endo, VLMC, Oligo, OPC, Micro/PVM). This procedure resulted in cell density estimates for each of the 24 cell-types at each cortical region using the same atlas as the synaptic density alteration maps. Due to data availability, non-cortical areas were excluded for analyses that used these cellular data.

Given that regional neurochemical and cellular profiles are multivariate, we examined enrichment between each of these two features and synaptic density alterations using robust non-parametric canonical correlation analyses^53^. Two canonical correlation analyses models were computed, examining the associations between regional synaptic density alterations and receptor (Fig. 2C) or cell densities (SFig5). Statistical significance of the observed canonical correlations (r_cca_) compared to null correlations derived using the spin-test^41,42^ with 10,000 permutations. Statistical inference was assessed at p<.05 (two-tailed). To examine which cell or receptor/transporter maps contributed most to synaptic density alterations, we quantified canonical loadings by computing product moment correlations between the 18 PET or 24 cell input maps and corresponding canonical variate. To assess statistical significance of canonical loadings, we used robust bootstrapping to estimate the standard error for each input map (10,000 bootstraps), computed z-scores by dividing the correlation by the standard error and used these z-scores to compute two-tailed p-values^54^. Maps showing reliable loadings on the canonical variate were those that survived a Bonferroni FWE-correction across input features (p_FWE_ < 0.05; two-tailed; Fig 2D).

### Network diffusion model

We used a network diffusion model to test whether the pattern of lower synaptic density is consistent with a diffusion process that spreads through the brain and the extent to which certain brain regions may act as sources, or initiation sites, of pathological spread (Fig4)^55^. The network diffusion model simulates the dynamic spread of pathology between the nodes of a weighted network via a process of diffusion (Fig4A), defined as

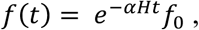

where 𝑡 is the model diffusion time, which has arbitrary units (a.u.), and 𝑓(𝑡) is a vector characterizing the amount of diffusion in each region at time 𝑡 . The model was constrained by a weighted structural connectivity matrix, as the base of diffusion modelling. Here, the structural connectivity matrix was derived using tractography applied to high-resolution diffusion-weighted imaging data the Human Connectome Project^56^ (details of the procedure provided in the Supplement). The strength of the diffusion process is controlled by a constant (𝛼) and 𝐻 is the Laplacian of the weighted base matrix. 𝑓_0_ represents the initial distribution of pathology. We repeatedly initialized the model using each of the 332 regions as the starting seed, such that the initial state was set to 1 for the seed region, and 0 for all other regions. At each initialization, using a constant of 𝛼 = 1, the network diffusion model was used to estimate the diffusion at all other regions at time 𝑡 = 0 to 50. In this way, we were able to determine whether a diffusion process seeded from each region resulted in a spatial distribution of synaptic density loss that matched the empirically observed patterns.

Consistent with prior work^20,55,57^, model performance was evaluated as the product-moment correlation between the predicted diffusion and observed volume abnormalities at each time step and for each seed, with the maximum correlation across all time steps. The seed region was excluded when correlating predicted and observed volume abnormalities to ensure that our analysis was not influenced by large synaptic abnormalities in the seeds. The performance of the model in capturing the empirical maps of synaptic density abnormalities was compared to its performance in capturing surrogate maps generated using the two benchmark models derived using either the spin-test applied to the empirical synaptic density alteration map, or a network rewiring algorithm applied to the empirical structural connectivity matrix. Seed regions were considered significant if they were assigned a p_FWE_ <.05 for both benchmark null models. Further details about benchmark null models used to evaluate the network diffusion model can be found in the Supplement.

## Results

### Widespread synaptic density abnormalities in schizophrenia

We find prominent and widespread pattern of lower synaptic density in individuals with schizophrenia (Fig. 1A), compared to controls, with frontal, temporal, cingulate, thalamic, striatal, hippocampal and cerebellar areas surviving stringent voxel-level family-wise error (FWE) correction (0.58 < Cohens *D* < 1.47; 𝑝_𝐹𝖶𝐸_ < 0.05, Fig. 1A). An analysis of hemispheric asymmetry indicated that the left hemisphere was substantially more impacted than the right hemisphere (*D* = 1.14, p < 0.001). We repeated this analyses after adjusting for antipsychotic medication exposure (SFig. 1A), after excluding individuals with a history of substance abuse (SFig. 1B), and after removing partial volume correction (SFig. 1C), finding a highly similar and significantly correlated pattern of lower synaptic density. Mean voxel-level *BP*_ND_ maps for patients and controls are provided in SFig. 2).

### Synaptic density abnormalities in schizophrenia do not overlap with grey matter abnormities

The brain-wide spatial patterns of synaptic and volumetric alteration were not correlated within the current (𝑟 = 0.03; 𝑝 = .0496), nor across four independent samples (Fig. 1C). This was not because the current sample was neuroanatomically distinctive, since there were significant correlations between the volume alteration maps of the current sample and those from other datasets (0. 33 < 𝑟 < 0.41; 𝑝_𝐹𝖶𝐸_ < 0.05), except for the first episode psychosis dataset (𝑟 =. −0.02; 𝑝 = 0.81). This is consistent a reduced magnitude of grey matter alterations reported in first episode psychosis, compared to chronic schizophrenia. These results remained consistent when examining each hemisphere independently (SFig. 3A), when deformation-based, rather than voxel-based, morphometry was used to measure gray matter volume differences between groups (SFig. 3B) and, when partial volume correction was not implemented for the synaptic density maps (SFig. 3C).

**Fig 3.**
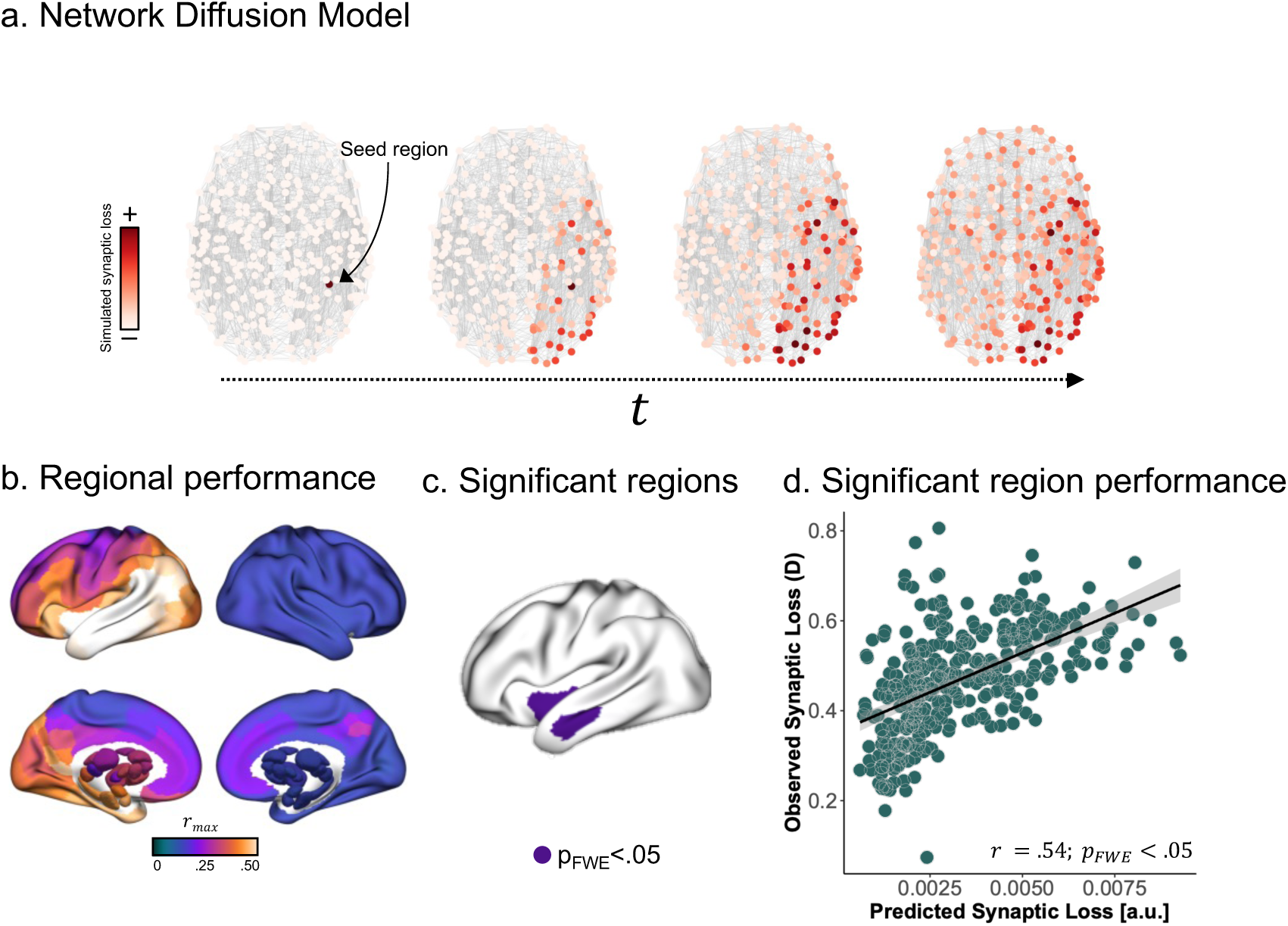
Network diffusion model of synaptic pathology. **(a)** To understand how global connectivity and local neurochemical vulnerabilities shape synaptic pathology, we simulated a spreading process using a network diffusion model. Using each of the 332 parcellated regions as a seed, we retained the maximum correlation between the simulated and observed synaptic abnormalities. We then compared maximum *r* values (r_max_) for each region to a distribution of maximum *r* values from two benchmark null models accounting for spatial autocorrelations in the synaptic pathology maps and basic topological properties of the connectivity matrix (see Methods). **(b)** Model performance (r_max_) for each region. **(c)** Seed regions (purple) with significantly (𝑝_𝐹𝖶𝐸_ < .05) greater r_max_ than both null models **(d)** Observed and network diffusion model predicted synaptic deficit, using the best performing seed (left frontal operculum/anterior insula).

Overall, these results demonstrate prominent and widespread lowering of synaptic density in schizophrenia, which is left lateralized and spatially independent of commonly reported MRI-derived gray matter volume alterations across different stages of illness.

### Network & molecular enrichment of synaptic pathology

Brain abnormalities in psychiatric and neurological disorders preferentially affect specific systems such as macroscale functional networks, distinct cytoarchitectonic regions and neurotransmitter systems^11,58,59^. This spatial organization of pathology suggests that the functional and molecular properties of distributed brain regions may confer differential vulnerability to synaptic pathology. Supporting this conjecture, in the right hemisphere, we find evidence for preferential accumulation of synaptic pathology within aspects of the functional frontoparietal/cognitive control network (FPN), in particular, dorsal precuneus and posterior cingulate areas (FPN C subdivision; 𝑍 = 3.02; 𝑝 < .001; Fig. 2A). We also find preliminary evidence that while the temporoparietal network is preferentially impacted in the left hemisphere (𝑍 = 1.70; 𝑝 = 0.047; Fig. 2B), it is relatively spared in the right (Z=-1.72; p = 0.025; Fig. 2A), suggestive of functional network-level asymmetry in synaptic pathology. No significant enrichment was observed for any cytoarchitectonic types of laminar differentiation.

We found a significant association between the spatial pattern of synaptic alterations and neurotransmitter concentrations within the left hemisphere (𝑟_𝑐𝑐𝑎_ = 0.68; 𝑝 = 0.022 ; Fig. 2C). When examining the canonical loadings of each receptor/transporter, we found that regions with typically higher concentrations of GABA_A/BZ_, 5HT_2A_, mGluR5 and 5HT_1B_, were reliably (𝑝_𝐹𝖶𝐸_ < 0.05; Fig. 2D) associated with lower synaptic density in individuals with schizophrenia. No significant neurotransmitter association was detected in the right hemisphere and no significant cell type associations were detected in either hemisphere.

### Synaptic alterations may be constrained by white-matter pathways

To understand the spatiotemporal dynamics of synaptic loss in schizophrenia and to identify regions from where pathology may initiate, we used a network diffusion model^55^ to directly test whether synaptic loss propagates through the brain in a way consistent with a process of diffusion between axonally connected regions (Fig. 3A). The performance (𝑟_𝑚𝑎𝑥_) for each brain region is shown in Fig. 3B. Two regions, both in the left hemisphere showed the best performance and surviving FWE correction (𝑝_𝐹𝖶𝐸_ < .05) across both null models: frontal operculum/anterior insula (𝑟_𝑚𝑎𝑥_ = 0.54; Fig. 3C) and the middle temporal gyrus (𝑟_𝑚𝑎𝑥_ = 0.53; Fig. 3C). Observed and predicted synaptic pathology using the best performing seed region (left frontal operculum/anterior insula) are shown in Fig 4D. These findings suggest that synaptic pathology may initiate in left hemisphere inferior frontal and middle temporal regions and spread to structurally connected regions.

Given the previous association found between neurotransmitter systems and synaptic alteration, we examined an exploratory network diffusion model where the interregional connectivity matrix was weighted by receptor/transporter similarity between regions (see Supplement for details). In this way, the model preferentially diffused pathology not only between regions that were structurally connected, but between those that also share a similar neurotransmitter composition. In this model, the left frontal operculum/anterior insula areas again showed the best performance survived FWE correction (𝑝_𝐹𝖶𝐸_ < .05) across both null models, suggesting that synaptic pathology may propagate between structurally connected and molecularly similar areas.

## Discussion

We show a brain-wide pattern of lower *in vivo* synaptic density in individuals with schizophrenia, compared to a large sample of healthy participants. This pattern is left lateralized and spatially independent of commonly reported MRI-derived gray matter volume alterations across different stages of psychotic illness. Functionally defined frontoparietal/control areas were enriched for synaptic pathology in the right hemisphere, whereas the spatial pattern of lower synaptic density left hemisphere is strongly coupled to the normative neurochemical composition, with areas rich in GABA_A/BZ_, 5HT_2A_, mGluR5 and 5HT_1B_ receptors most strongly affected. Using simulation-based network diffusion models, left inferior frontal/anterior insula areas were identified as initial sources from which synaptic pathology initiates and spreads to axonally connected and neurochemically similar areas.

### Widespread lower synaptic density in schizophrenias

The quantification of synaptic density within the living brain used [^11^C]UCB-J, a PET tracer that targets the Synaptic Vesicle Protein 2A (SV2A)^6,60^ which is localized and monodispersed on vesicular membranes within all chemical pre-synaptic boutons ^61–63^. This tracer has highly specific binding to SV2A^24^, good test-retest reliability^6,64^, and displacement within gray matter using a drug with specific binding to SV2A^6^. In primates, regional SV2A levels are strongly correlated (r >.95) with synaptophysin^6^, a validated marker of *ex vivo* presynaptic density. Moreover, [^11^C]UCB-J uptake recapitulates anatomically inferred patterns of synaptic density^6^, has been shown to be sensitive to synaptic pathology across multiple neurodegenerative conditions^6,65–67^, and is specific to synaptic loss measured using electron microscopy, rather than reductions in protein levels^68^. Nonetheless, it is possible that the alterations detected here represent reduced SV2A or vesicular organelles in schizophrenia, in the absence of changes in terminal numbers. For instance, initial evidence suggested that a polymorphism in the SV2A gene may increase risk for schizophrenia, though this has not been replicated in recent large-scale genome-wise association studies^69^. Furthermore, while sustained vesicular depletion occurs as a secondary effect in neurological disorders initiated by illness-linked proteins^70–72^, limited post-mortem evidence in schizophrenia actually suggests increased numbers and clustering of synaptic vesicles^73,74^—findings that may result from *ex vivo* artifacts^75,76^. Given the sensitivity of [^11^C]UCB-J to detecting synaptic terminal loss in other neurological conditions, and the strong converging genetic and post-mortem evidence demonstrating lower presynaptic terminals and dendritic spines in schizophrenia, it is likely that lower levels of [^11^C]UCB-J binding detected here in schizophrenia are likely a result of lower numbers of synaptic terminals.

We find that areas of gray matter in individuals with schizophrenia, especially thalamic, striatal, hippocampal, frontal, temporal, cingulate, cerebellar and occipital regions, show lower levels of synaptic density compared to controls, with medium to large effect sizes (0.58 < 𝑑 < 1.47). These findings align with previous region-of-interest studies examining *in vivo* synaptic density in early psychosis and schizophrenia populations, which have reported lower tracer binding in frontal, anterior cingulate, hippocampal, temporal, occipital and striatal regions^7–9,77,78^, using both measures of specific binding to SV2A, such as that used in the current study, and measures that additionally index non-specific binding. Extending this prior work, our results suggest that synaptic alterations are not only isolated to these regions but are far more widespread and left lateralized in chronic schizophrenia than previously reported. Our finding are also consistent with large-scale neuroanatomical studies in schizophrenia showing subtle left lateralized cortical thinning driven by regions of the language network ^79^.

Patients with schizophrenia often exhibit alterations in cortical thickness and gray matter volume^80–85^, which can progressively worsen in some individuals^37,86,87^. Given the limited evidence from post-mortem studies for a reduction in the number of neurons, it has been theorized that the substrate underlying these MRI abnormities are alterations in synaptic, dendritic, and axonal organization^88^. However, there is limited direct evidence linking MRI signals to neurite changes in psychotic illnesses. Alternatively, MRI signal differences between patients and controls may reflect differences in properties related to myelination, soma, glia, vasculature, or extracellular space. The lack of neurobiological understanding behind MRI signal alterations in psychotic illness have led others to suggest that artefactual alterations between cases and controls in hydration, cholesterol levels, physical and mental activity, and stress^89–91^ may be causes of the observed volumetric changes. Our finding that the pattern of synaptic and gray matter alterations does not spatially correlate within the same or independent samples across different stages of illness suggests that a lowering of presynaptic terminals is not the primary substrate underlying gray matter MRI abnormalities in psychotic illness. This finding is also consistent with prior work demonstrating that age-related loss of synaptic density has a different spatial pattern to age-related MRI-derived gray matter volume loss^92^. Although, future work should examine association with a broader range of MRI-derived gray matter metrics such as cortical thickness and surface area.

### Local molecular vulnerability and global axonal connectivity constrain synaptic abnormalities

In many neurological illnesses, pathological processes selectively target or initiate from specific genetic, cellular^93,94^, neurochemical^95,96^, and functional systems^10,97^. Here, we find evidence for the preferential accumulation of synaptic pathology within functionally defined control systems, specifically dorsal precuneus and posterior cingulate areas. This is consistent with a large body of work demonstrating that higher-order transmodal association areas are a primary site of pathology in psychosis^98,99^. Our findings also indicate a strong spatial correspondence of left hemisphere synaptic pathology with normative receptor profiles. Specifically, regions typically rich in GABA_A/BZ_, 5HT_2A_, mGluR5 and 5HT_1B,_ receptors, show vulnerability to synaptic pathology. All three neurotransmiter systems have been shown to be disrupted or track symptom severity in schizophrenia^100–102^, suggesting that synapses associated with these systems are vulnerable to pathology. This is also consistent with genetic findings showing that enrichment of common variant associations is restricted to genes that are expressed in both excitatory and inhibitory neurons^103,104^ and in genes encoding proteins involved in general synaptic features. Notably, we did not find associations with dopamine receptors or transporters, despite finding lower synaptic density in striatal regions, likely because our brain-wide spatial analysis is dominated by variation in cortical patterns that overlaps poorly with the predominantly striatal distribution of dopamine maps. Moreover, dopaminergic abnormalities in schizophrenia are thought to reflect presynaptic functional dysregulation rather than regional receptor/synapse loss.

We also did not find that densities of transcriptomically-defined cell types were predictive of the spatial pattern of synaptic pathology. Many post-mortem and genetic studies have shown layer specific cell abnormalities, commonly in frontal supragranular cells^105,106^ and particularly in cortical layer three^107–109^.

The current spatial resolution of PET does not allow for layer specific inference, therefore a spatial correspondence with layer specific cell types may become apparent with improving resolutions.

We used a network diffusion model to understand the spatiotemporal dynamics underlying the local molecular and brain-wide connectivity constraints. We find that left frontal operculum/anterior insula and middle temporal cortices are a putative initial source of synaptic density loss in schizophrenia. Neuroanatomical alterations in these regions are some of the earliest neural alterations reported in the initial stages of illness^110–112^. Our exploratory analysis which parametrized the network diffusion model using a neurotransmitter similarity matrix again nominated left frontal operculum/anterior insula areas as putative source regions. This is consistent with recent large-scale studies that have subtyped schizophrenia based on the trajectory of gray matter changes, which found that the most common subtype is characterized by initiation of pathology in inferior fronto-insular areas^21,113^. Our results are in line with a spreading process in which pathological processes preferentially propagate via axonal connections to molecularly similar areas. The precise mechanism driving this process remains unclear. Trans-neuronal spreading processes have often been seen in prion-like spread of misfolded proteins, but limited evidence exists for visible deposits of aggregated pathological proteins in psychotic illness (although see^114–116^). However, although speculative, subtle changes in protein homeostasis^117^ may interact with regional neurotransmitter levels^118,119^ and spread to synaptically connected distal brain region^120,121^. Alternatively, and given the commonly reported finding of functional brain alterations in psychotic disorders, dysfunction of one region may trigger abnormal functional activity in connected sites that, over time, may trigger synaptic changes as a result of aberrant neurotransmission or a loss of trophic support^122^. Indeed, trans-neuronal propagation of excitatory-inhibitory imbalance, with consequent excitotoxic stress, could plausibly contribute to the reduced synaptic density observed in schizophrenia^123^.

### Limitations

Our analysis of synaptic density depended on group-level summary values and may not directly reflect synaptic differences for individual patients. While the extent of interpatient heterogeneity in synaptic pathology is currently unknown, subsequent work could use larger samples with synaptic density imaging to derive patient-specific measures, such as those obtained through normative modelling^124,125^. The cross-sectional nature of this study precludes inference about the temporal onset of synaptic pathology, and further longitudinal studies will be able to clarify whether these alterations occur during neurodevelopment, at transition to illness or at later stages. An important consideration here is that the cell and receptor density maps used in the current study were derived from healthy samples and therefore represent normative cytoarchitecture and chemoarchitecture. Future data aggregation efforts examining cell-type and receptor maps from patient populations may provide additional evidence regarding synaptic vulnerabilities in schizophrenia. Our approach to characterize structural connectivity is also limited by the accuracy of diffusion MRI^126^. While the processing procedures we applied enhanced the biological validity of our structural connectivity measures as much as possible^127^, further developments in non-invasive connectivity mapping and tractography will be required to allow more precise mechanistic inferences about constrains imposed by axonal pathways.

Overall, our findings provide *in vivo* evidence for widespread presynaptic terminal deficits in schizophrenia that are left-lateralized, independent of gray matter alterations, aligned to specific neurochemical systems, and suggest that such synaptic pathology may propagate in a way consistent with axonal networks.

## Data Availability

All data produced in the present study may be based available upon request to the authors and ethical approval form appropriate an IRB.

## Acknowledgements

SC is supported by the University of Melbourne McKenzie Fellowship and the Brain Behavior Research Foundation. IE (Nancy Taylor Foundation and VA NCPTSD), DM (NINDS R01NS124819)). AS is supported by the Yale Wu Tsai Postdoctoral Fellowship). VC is supported by an Australian NHMRC Investigator Grant (1177370) and a University of Melbourne Dame Kate Campbell Fellowship. This study was supported in part by the National Institute on Drug Abuse (NIDA) grant R01 DA052454-03 (GAA), U54 AA027989 (KC), Dana Foundation David Mahoney program (RR), CTSA Grant Number UL1 TR001863 from the National Center for Advancing Translational Science (NCATS) (RR), National Center for Homelessness among Veterans (36C24820Q1276) (RR), and National Institute for Mental Health (NIMH) grant R01MH120080 (AJH). We also thank Dr. Jack Tsai (NCHAV) for administrative support.

