## Supplement for "Network-based Molecular Constraints on *in vivo* Synaptic Density Alterations in Schizophrenia"

***Normative reference structural connectome construction***

To estimate inter-regional axonal connectivity, we computed a representative weighted connectivity matrix from tractography applied to diffusion-weighted imaging (DWI) data from an independent healthy unrelated cohort of 150 (71 males, aged 21-35 years) individuals from the S900 release of the Human Connectome Project[1], which served as a reference connectome for computational modelling. This subset of 150 individuals was selected to reduce computational burden, and represented individuals with the lowest head motion, as estimated using framewise displacement[2]. Data were acquired on a customized Siemens 3T Connectome Skyra scanner at Washington University in St. Louis, MO, USA, using a multishell protocol for the diffusion weighted imaging with the following parameters: 1.25-$mm^3$ voxel size; repetition time = 5520 ms; echo time = 89.5 ms; field of view of 210 mm by 180 mm; 270 directions with $b$ = 1000, 2000, 3000 s/$mm^2$ (90 per $b$ value); and 18 $b$ = 0 volumes. Structural T1-weighted data were acquired with 0.7 mm voxels, repetition time = 2400 ms, echo time = 2.14 ms, and a field of view of 224 $mm^2$.

The DWI data first underwent the HCP minimal pre-processing pipeline[3], that included normalization of mean $b$ = 0 images across diffusion acquisitions and correction for echo-planar imaging susceptibility and signal outliers, eddy current–induced distortions, slice dropouts, gradient nonlinearities, and participant motion. T1-weighted data were corrected for gradient and readout distortions before being processed with FreeSurfer[4]. Further details regarding this pipeline are provided elsewhere[5]. Using the corrected DWI data, fiber orientation distributions were estimated using multishell Constrained Spherical Deconvolution[6]. Probabilistic tracking was performed with the Fiber Orientation Distributions (iFOD2) algorithm, implemented using MRtrix3[7,8] and used Anatomically Constrained Tractography which takes advantage of previously computed tissue maps to ensure that streamline can begin, traverse, and terminate in anatomically plausible locations[9]. A total of 10 million streamlines were generated using dynamic seeding along with default MRtrix3 parameters[7,10].

The same 332 regions atlas (the homotopic version was not available at time of connectome construction[11,12]) was then transformed from template surface space to the individual surface of each subject using the spherical registration[13]. The same atlas was used to parcellate the synaptic density alteration map for the network diffusion model. Next, the parcellation was projected to a volumetric image and resampled to the same resolution as the DWI using nearest-neighbor interpolation[14]. To create a connectivity matrix, streamlines were assigned to each of the closest regions in the parcellation within a 5mm radius of the streamline endpoints[7].

Importantly, most tractography algorithms are prone to false positives and do not directly index the quantitative strength of connections between pairs of regions[15,16]. We therefore implemented a state-of-the-art optimization procedure, Convex Optimization Modelling for Microstructure Informed Tractography (COMMIT2), which has shown to be superior to other methods on key benchmarks derived from fiber-tracking phantoms[17]. COMMIT2 uses a forward model to recover the connectivity matrix with the minimum number of bundles that best explains the local axon density estimated from the DWI signal[17]. In doing so, the algorithm filters and re-weights inter-regional connectivity strengths and provides more biologically accurate quantitative estimates of connectivity. After optimized connectivity matrices were generated for each subject, we computed a single group-consensus matrix by retaining connections if they appeared in at least $\tau$ subjects, where $\tau$ is the consensus threshold that results in a binary density comparable to that of a typical subject[18], and which was set to 37.7% for this sample. This threshold is computed separately for inter-/intra-hemispheric connections. Retained connections are assigned the corresponding group-average connectivity weight, resulting in a weighted consensus connectivity matrix. Finally, the connectivity weights from the group- consensus matrix were z-scored. This procedure resulted in a single 332 × 332 weighted connectivity matrix.

***Benchmark null model details for network diffusion model***

The network diffusion model tests the spreading hypotheses by simulating a passive diffusion process to model the spread of synaptic density alterations from specific seed, or 'epicentre', regions from which the spread may initiate. We seeded the network diffusion model using each of the 332 individual brain regions as a seed region for each seed region from the parcellated synaptic density alteration map (Fig1A). For each simulation, the measure of accuracy used to evaluate the network diffusion model was the maximum correlation obtained across different time steps of the diffusion process ($r_{max}$; Fig3B) between the simulated and observed synaptic density alteration. Note that, in principle, the network diffusion model could be initialized using any combination of seed regions[19,20], but this can quickly result in a combinatorial problem of alternative initial conditions for the model, therefore we focused on testing specific hypotheses about individual brain regions as putative sources of synaptic density alteration.

To evaluate the statistical significance of each region's $r_{max}$, we generated a null distribution of $r_{max}$ values using two benchmark models: the $\text{Null}_{\text{spin}}$ and the $\text{Null}_{\text{rewire}}$ null. The $\text{Null}_{\text{spin}}$ model evaluated whether the observed findings were specific to the empirically observed pattern of synaptic density abnormality or were a generic property of the intrinsic spatial structure of the abnormality map. The model uses a spin-test to rotate region-level cortical test-statistics 1000 times[21], with subcortical regions randomly permuted without replacement. The rotation was applied to one hemisphere and then mirrored for the other hemisphere. This is the same benchmark used for all other analyses in the paper. The second null model ($\text{Null}_{\text{rewire}}$) involved rewiring the structural connectome while preserving the degree sequence and length-weight relationship, and approximately preserving the edge-length distribution[22]. As per previous work[23], we used 10 distance bins and 50,000 edge swaps to generate 1000 rewired networks. These surrogate networks were used to test the hypothesis that any apparent network-based prediction of local synaptic denisity is specific to the actual topology of the connectome itself, and cannot be explained by basic network properties, such as regional variations in node degree or the spatial dependence of inter-regional connectivity.

To generate null $r_{max}$ values for each contrast using the $\text{Null}_{\text{spin}}$, we initiated the network diffusion model from each brain region 1000 times and evaluated the correlation between the simulated synaptic density loss and the spatially constrained null synaptic density loss generated by the $\text{Null}_{\text{spin}}$. The $r_{max}$ from each iteration was retained, resulting in 1000 null $r_{max}$ values at each brain region. For each contrast and each brain region, the p-value was considered as the percentage of nulls $r_{max}$ null values greater than the observed $r_{max}$. To implement FWE correction, the maximum brain-wide null $r_{max}$ from each of the 1000 iterations was used to construct a FWE-corrected null distribution[24]. The FWE-corrected p-value was considered as the percentage of FWE-corrected null $r_{max}$ values greater than the observed $r_{max}$. To evaluate significance using the $\text{Null}_{\text{rewire}}$ null model, we followed the same procedure described above, but instead of varying the synaptic density loss, we varied the structural connectome at each of the 1000 iterations, using null connectomes generated using the rewiring method. Regions were considered significant seeds if they passed the $p<.05$ FWE threshold for both $\text{Null}_{\text{spin}}$ and $\text{Null}_{\text{rewire}}$.

***Normative reference receptor/transporter similarity matrix construction***

For regional normative neurochemical densities, we used 18 different group-consensus PET binding maps from healthy population collated by the *neuromaps* toolbox[25]. Following previous work, for tracers that had multiple samples, a sample-size weighted average map was computed[26]. For cases where two different tracers targeted the same receptor/transporter, the one with the larger sample size was retained. Further details regarding each PET map are provided elsewhere[25]. Each PET map was masked and parcellated into the same 332 region atlas as the synaptic density alteration maps used for the network diffusion modelling. Inter-regional receptor/transporter similarity were estimated using the same parcellated 18 PET binding maps. Regional binding values were first z-scored within each map and pair-wise product moment correlations were computed between each region's receptor/transporter profile, followed by a Fishers z-transformation. Finally, the matrix was min-max transformed to scale the similarity values between [0,1] as the network diffusion model cannot account for negative weights.

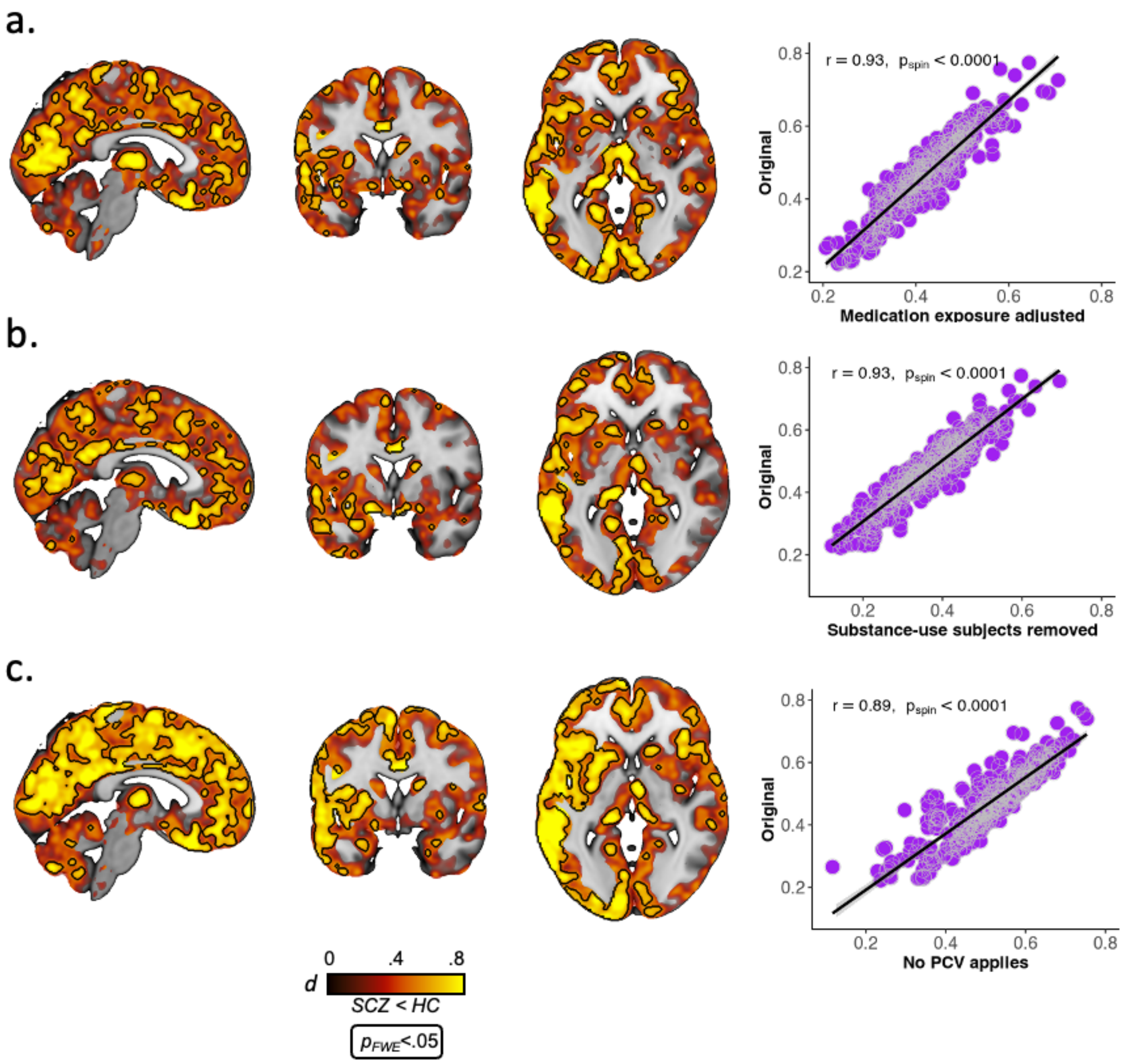


**SFig. 1 –** Effect size maps for group difference in synaptic density after adjusting for medication exposure **(a)**, removal of subjects with substance abuse **(b)**, removing partial volume correction (**c**; PVC). Black outline shows voxel-level FWE-corrected significant areas. Scatter plots show association between each parcellated effect size map and the group difference maps used in the primary analysis (Fig. 1A).

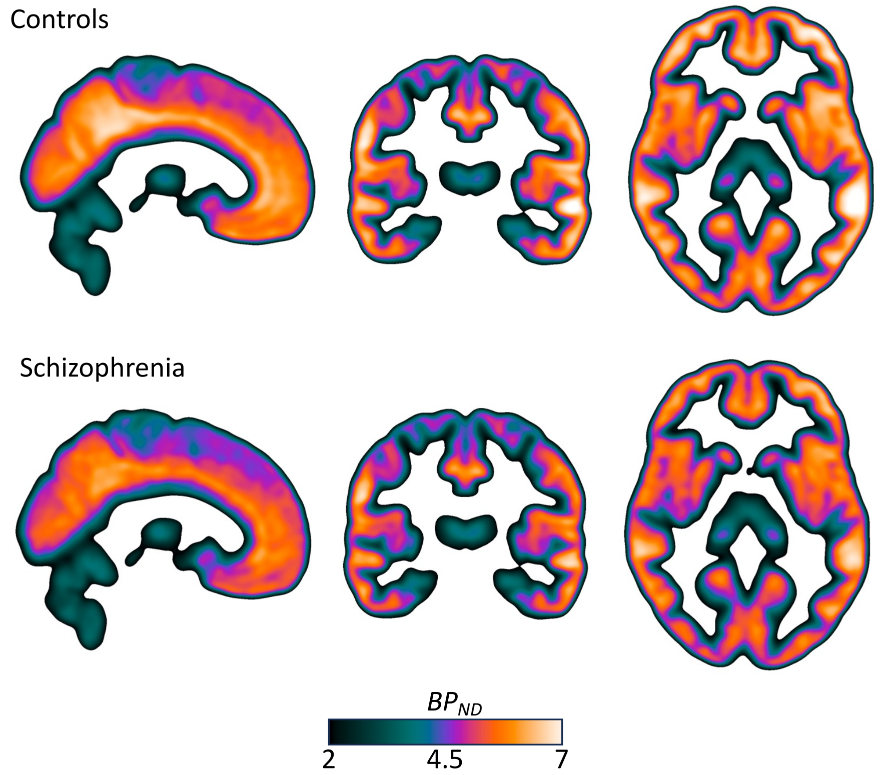


**SFig. 2 –** Mean of parametric $BP_{\mathrm{ND}}$ images for controls (top) and patients (bottom). Linear interpolation has been applied to the images for visualization.

a. Associations with GMV (split by hemisphere)

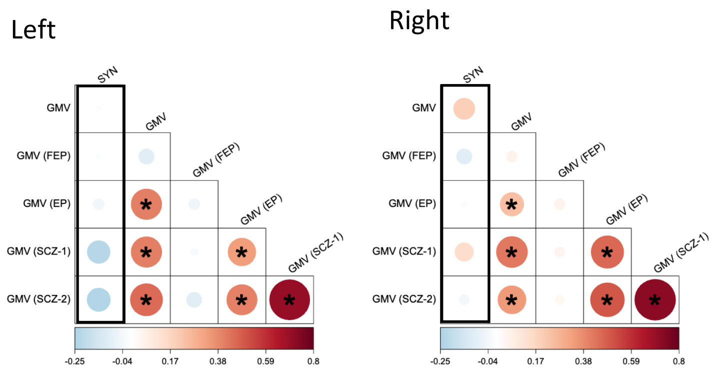


b. Associations with GMV (using DBM)

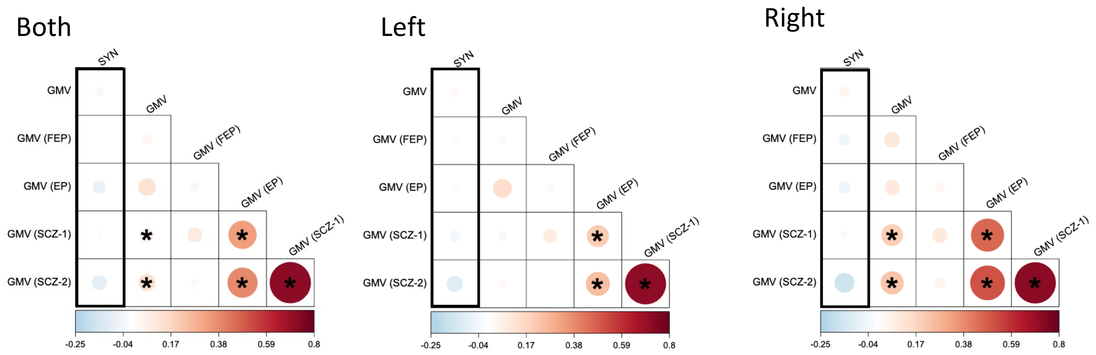


c. Associations with GMV (without PVC on PET data)

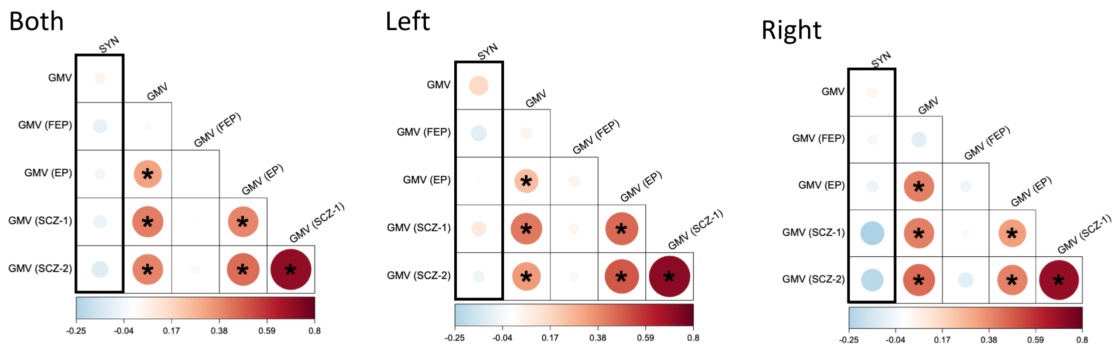


**SFig. 3 –** Spatial correlation (Pearsons r) between parcellated synaptic density alterations (SYN) and gray matter volume (GMV) within the current sample (top left box in each panel), as well as four independent samples including antipsychotic-naive first-episode psychosis (FEP), early psychosis (EP) and two established schizophrenia (SCZ) cohorts. All other boxes represent correlations between GMV maps between the current and independent samples. **(a)** Correlations when hemispheres are examined separately, when **(b)** synaptic density data is not partial volume corrected (PVC) and, **(c)** when DBM is used to measure grey matter alterations, instead of VBM.
